# COVID-19 Trends in Florida K-12 Schools, August 10 – November 14, 2020

**DOI:** 10.1101/2020.11.30.20241224

**Authors:** Rebekah D. Jones

**Author notes:** **Address correspondence to**: Rebekah Jones, The Covid Monitor, PO BOX 13747, Tallahassee, FL 32317. **Conflict of Interest Disclosures (includes financial disclosures):** The author has no conflicts of interest to disclose. **Funding/Support:** No funding was secured for this study.

## Abstract

Data collected from 38 states from August 3 – November 15, 2020 showed more than 250,000 confirmed student and staff cases of SARS-CoV-2 in K-12 schools^1^. Yet, analysis of COVID-19 case data in USA schools has been extremely limited^2,3^. To date, no large-scale or state-wide analyses by school level and grade has been published, opening a wide gap in understanding COVID-19 in American schools. A large-scale assessment of available data and trends could provide a baseline for understanding the virus in the K-12 learning environment and dispel misconceptions about the prevalence of COVID-19 in schools.

**Table of Contents Summary:** Using the most comprehensive database of K-12 COVID-19 case data in the country, Florida provides clues for understanding student and staff cases in schools.

*What’s known on this subject:* Florida schools began reopening to in-person instruction in August have reported more than 18,000 student and staff cases of COVID-19 as of November 14, 2020. Incidence of COVID-19 cases in K-12 students and staff is of urgent public health concern.

*What this study adds:* COVID-19 cases reported in Florida schools were most influenced by community case rates, district mask policies, and percent of students attending face-to-face. Student case rates were highest in high schools (12.5 per 1,000).

## Background

Florida school case data provides an opportunity to examine the extent to which COVID-19 has been detected and reported in schools through reporting of cases by day, by school level, by location, for both students and staff. Student enrollment across Florida’s 67 districts totals more than 2.67 million, with five of the ten most populated districts in the country within Florida’s public-school system. Schools in Florida’s mix of urban, suburban and rural districts, in addition to the variety of policies ranging from availability of virtual instruction to mandatory-mask mandates, make it an ideal case study for examining larger trends in COVID-19’s prevalence in American schools. This data could help inform decisions makers evaluating mitigation strategies and access to virtual learning, as well as build upon current knowledge of COVID-19 in American society.

## Methods

During August 10 – November 14, 2020, laboratory-confirmed cases of COVID-19 in 10,088 students and 4,507 staff in K-12 schools were confirmed by either the Florida Department of Health, or directly from the 43 independently-reporting school districts in the state (Table 1). School case data, collected daily, and school enrollment data for both in-person (including hybrid) and virtual learning were obtained by public records request to each of Florida’s 67 districts for each week of the study period. Data regarding mask policies were obtained either from each district’s reopening plan, or from public records request. No assessment of school case data or rates with consideration of age groups and mask policies at this scale or with this level of granularity were discovered after an exhaustive search.

**Table 1:**
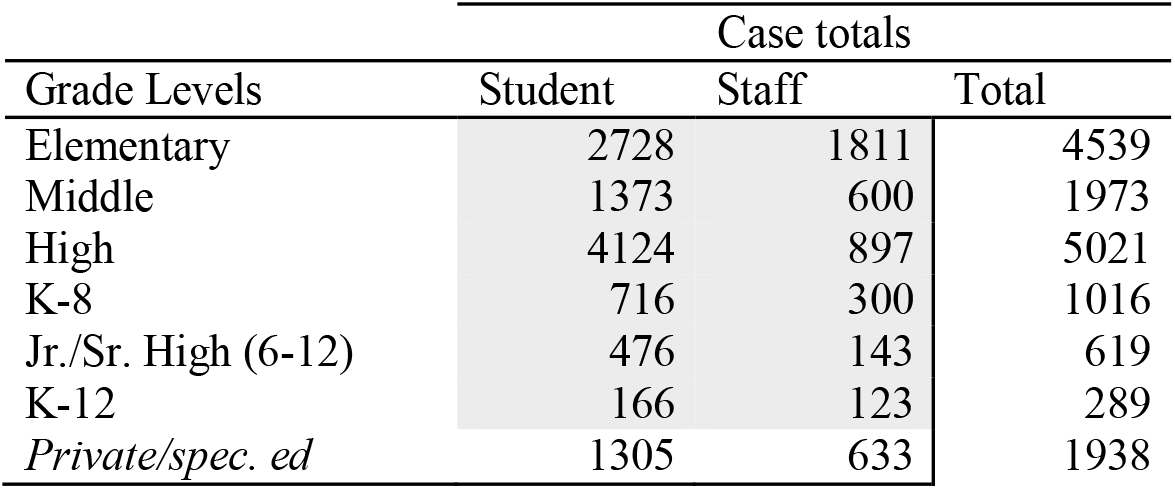
Florida COVID-19 case totals by school level for students and staff, August 10—November 14, 2020.

| Grade Levels | Case totals |  |  |
| --- | --- | --- | --- |
|  | Student | Staff | Total |
| Elementary | 2728 | 1811 | 4539 |
| Middle | 1373 | 600 | 1973 |
| High | 4124 | 897 | 5021 |
| K-8 | 716 | 300 | 1016 |
| Jr./Sr. High (6-12) | 476 | 143 | 619 |
| K-12 | 166 | 123 | 289 |
| *Private/spec. ed* | 1305 | 633 | 1938 |

The Centers for Disease Control (CDC) provides guidance for safely reopening schools, based primarily on the rate of community spread within a given district or county^5^. County COVID-19 case rates were calculated based on new cases over a 14-day period at a rate per 1,000 people based on data reported by the Florida Department of Health^6^.

## Results

During August 10 – November 14, 2020, the state-wide incidence rate (cases per 1,000 students enrolled in face-to-face or hybrid instruction) in Florida high school students (12.5) was 70% higher than younger cohorts (7.4). School data by grade level (e.g. elementary, middle, high) shows trends consistent with earlier findings by the CDC regarding case rates between younger and older adolescents^4^. Staff rates are higher than student rates in all school environments except high schools. The proportion of student to staff cases in Florida schools was closest in the elementary setting (60% students – 40% staff) compared to the high school setting (82% students – 18% staff).

Approximately 61% of all students in Florida returned to in-person instruction during the report period, with 39% enrolled in virtual-learning programs or withdrawing from the district by November 1, 2020. Most students (87%) in the state who attended in-person classes were enrolled in districts with mandatory mask mandates, though the percent of students enrolled in face-to-face instruction was highest among districts without mask mandates. The state-wide case incidence rate in districts without mask mandates (12.7 per 1,000 students enrolled face-to-face or hybrid) was 38% higher than those who attended in-person within districts that did have mask mandates (9.2). However, the staff case rate in districts without mask mandates (29.2 per 1,000 on-campus employees) was nearly twice that of staff case rates in districts with mandatory mask policies (14.8) (Table 2).

**Table 2:** Florida COVID-19 school-based case rates (August 10 – November 14, 2020), community and school-age case rates (November 1 – 14, 2020), and mask policies by county

| County | School Case Rates (per 1,000 students enrolled) |  |  |  |  | Community 14-day case data per 1,000 |  |  | Mandatory Mask Policy |
| --- | --- | --- | --- | --- | --- | --- | --- | --- | --- |
|  | Elementary Student | High Student | District Rate (all levels – all types) | District student rate (all levels) | District Staff Rate (all levels) | All age groups | School-age children (4-17) | Percent of school-age cases also reported by school district |  |
| Alachua | 8.2 | 26.1 | 12.1 | 10.0 | 23.6 | 3.6 | 13.0 | 48.5 | Yes |
| Baker | 6.7 | 34.6 | 14.0 | 11.3 | 34.2 | 3.2 | 3.5 | 45.5 | No |
| Bay | 6.6 | 16.3 | 9.6 | 8.0 | 20.7 | 3.4 | 18.9 | 75.0 | Yes |
| Bradford | 10.4 | 17.2 | 11.7 | 8.0 | 29.9 | 2.0 | 0.2 | 0.0 | No |
| Brevard | 6.8 | 9.1 | 6.8 | 6.6 | 14.3 | 2.9 | 16.6 | 55.2 | Yes |
| Broward | 5.1 | 3.3 | 4.7 | 3.6 | 10.4 | 3.6 | 182.2 | 36.0 | Yes |
| Calhoun | 6.1 |  | 12.2 | 19.2 | 30.4 | 4.2 | 3.9 | 55.6 | No |
| Charlotte | 3.0 | 7.3 | 5.3 | 5.2 | 5.8 | 2.8 | 2.5 | 66.7 | No |
| Citrus | 7.3 | 15.8 | 10.2 | 7.6 | 25.4 | 3.8 | 19.2 | 100.0 | Yes |
| Clay | 4.8 | 9.0 | 6.5 | 6.8 | 14.7 | 3.0 | 21.0 | 55.0 | Yes |
| Collier | 8.5 | 12.0 | 7.8 | 6.5 | 14.2 | 3.2 | 57.5 | 66.3 | Yes |
| Columbia | 7.2 | 10.6 | 10.9 | 11.4 | 24.7 | 2.9 | 11.4 | 85.7 | No |
| Desoto | 8.5 | 6.7 | 6.4 | 4.9 | 13.7 | 2.3 | 1.8 | 200.0 | No |
| Dixie | 1.9 | 22.8 | 6.1 | 4.6 | 15.0 | 1.4 | 0.1 | 50.0 | No |
| Duval | 4.8 | 9.9 | 6.8 | 5.8 | 14.9 | 2.7 | 25.7 | 35.3 | Yes |
| Escambia | 4.2 | 15.6 | 6.5 | 5.2 | 12.1 | 4.1 | 37.0 | 55.6 | Yes |
| Flagler | 6.3 | 11.6 | 8.2 | 7.6 | 16.9 | 1.8 | 3.5 | 95.7 | No |
| Franklin |  |  | 32.4 | 23.7 | 85.6 | 2.0 | 6.0 | 0.0 | No |
| Gadsden | 14.6 | 12.9 | 11.5 | 12.5 | 11.3 | 1.1 | 2.0 | 9.1 | Yes |
| Gilchrist | 3.7 |  | 10.0 | 16.8 | 29.3 | 2.0 | 8.9 | 170.0 | No |
| Glades | 31.4 |  | 14.2 | 13.1 | 68.8 | 1.6 | 0.5 | 200.0 | No |
| Gulf | 4.5 |  | 15.1 | 28.1 | 21.4 | 3.6 | 0.0 | 100.0 | No |
| Hamilton | 2.4 |  | 3.2 | 2.3 | 19.3 | 1.8 | 0.1 | 50.0 | No |
| Hardee | 22.2 | 50.1 | 29.5 | 30.8 | 53.3 | 2.2 | 1.1 |  | No |
| Hendry | 9.9 | 8.2 | 8.8 | 6.7 | 19.9 | 2.4 | 7.3 | 50.0 | Yes |
| Hernando | 3.5 | 8.4 | 6.6 | 5.7 | 11.7 | 2.2 | 14.0 | 48.5 | No |
| Highlands | 8.2 | 11.1 | 11.2 | 10.1 | 26.8 | 4.5 | 6.4 | 75.0 | No |
| Hillsborough | 8.6 | 14.0 | 8.5 | 6.0 | 20.5 | 2.7 | 38.9 | 75.3 | Yes |
| Holmes | 12.3 | 21.1 | 14.7 | 11.7 | 51.1 | 4.9 | 2.1 | 57.1 | No |
| Indian River | 8.4 | 18.6 | 12.1 | 11.5 | 14.5 | 3.1 | 2.6 | 76.4 | Yes |
| Jackson | 12.4 | 31.0 | 16.7 | 16.7 | 44.1 | 7.1 | 2.2 | 58.3 | No |
| Jefferson | 4.7 | 20.2 | 24.1 | 37.6 | 21.4 | 2.5 | 0.1 | 16.7 | Yes |
| Lafayette | 3.7 |  | 6.1 | 6.9 | 32.1 | 1.0 | 0.0 |  | No |
| Lake | 7.6 | 9.7 | 7.8 | 6.1 | 16.9 | 2.4 | 5.6 | 34.4 | Yes |
| Lee | 6.6 | 11.6 | 7.9 | 6.9 | 13.4 | 3.0 | 4.4 | 75.5 | Yes |
| Leon | 9.7 | 14.2 | 10.6 | 9.4 | 14.6 | 3.5 | 1.2 | 37.5 | Yes |
| Levy | 6.6 |  | 10.5 | 17.7 | 6.3 | 2.1 | 0.2 | 136.8 | No |
| Liberty |  | 22.6 | 17.7 | 17.0 | 21.6 | 1.7 | 0.0 |  | No |
| Madison | 5.9 | 13.3 | 11.1 | 9.5 | 26.6 | 4.8 | 1.8 | 55.6 | No |
| Manatee | 7.1 | 9.4 | 6.0 | 5.0 | 10.6 | 2.6 | 15.0 | 44.5 | Yes |
| Marion | 6.0 | 12.3 | 7.1 | 5.8 | 13.3 | 2.0 | 0.3 | 39.1 | No |
| Martin | 3.3 | 14.1 | 8.1 | 8.0 | 8.7 | 1.7 | 1.5 | 73.9 | Yes |
| Monroe | 5.7 | 8.2 | 5.0 | 5.3 | 6.6 | 5.4 | 25.5 | 83.3 | Yes |
| Nassau | 6.0 | 8.8 | 8.8 | 9.8 | 18.1 | 2.7 | 0.6 | 66.7 | Yes |
| Okaloosa | 9.6 | 45.0 | 18.1 | 15.6 | 38.8 | 4.8 | 0.9 | 77.5 | No |
| Okeechobee | 7.6 | 12.4 | 8.4 | 8.4 | 8.5 | 2.4 | 0.4 | 75.0 | Yes |
| Orange | 8.9 | 11.6 | 7.9 | 7.6 | 8.8 | 3.1 | 10.1 | 45.0 | Yes |
| Osceola | 17.1 | 14.1 | 13.8 | 13.3 | 15.9 | 4.0 | 11.4 | 43.3 | Yes |
| Palm Beach | 7.5 | 15.7 | 8.9 | 8.7 | 10.1 | 3.3 | 59.7 | 56.3 | Yes |
| Pasco | 8.6 | 14.8 | 9.1 | 8.5 | 12.9 | 2.6 | 18.1 | 83.7 | Yes |
| Pinellas | 6.4 | 12.4 | 9.7 | 9.0 | 12.5 | 2.8 | 42.2 | 61.1 | Yes |
| Polk | 7.6 | 8.4 | 7.0 | 6.5 | 11.4 | 2.5 | 0.9 | 42.4 | Yes |
| Putnam | 9.1 | 19.6 | 14.6 | 12.3 | 32.6 | 1.6 | 0.4 | 66.7 | Yes |
| Santa Rosa | 7.2 | 13.7 | 8.5 | 6.7 | 25.9 | 3.2 | 0.7 | 32.8 | Yes |
| Sarasota | 7.7 | 18.2 | 8.1 | 7.9 | 9.2 | 2.0 | 0.9 | 315.2 | Yes |
| Seminole | 6.5 | 29.8 | 9.9 | 9.9 | 9.9 | 2.9 | 0.7 | 151.6 | Yes |
| St. Johns | 6.7 | 13.8 | 8.7 | 7.9 | 14.3 | 2.9 | 1.0 | 115.5 | Yes |
| St. Lucie | 2.5 | 10.2 | 6.2 | 6.0 | 8.1 | 2.0 | 11.0 | 33.3 | Yes |
| Sumter | 8.9 | 14.2 | 11.5 | 15.7 | 20.0 | 1.5 | 0.2 | 100.0 | Yes |
| Suwannee | 10.1 | 16.1 | 14.1 | 11.7 | 28.9 | 2.1 | 0.3 | 76.9 | No |
| Taylor | 12.8 | 32.2 | 13.7 | 12.8 | 18.0 | 3.1 | 0.2 | 72.7 | No |
| Union | 12.7 | 66.0 | 21.9 | 19.1 | 41.2 | 2.9 | 0.2 | 87.5 | No |
| Volusia | 5.5 | 11.4 | 7.6 | 6.4 | 14.6 | 2.2 | 3.1 | 58.1 | Yes |
| Wakulla | 11.3 | 18.9 | 11.1 | 10.6 | 14.0 | 3.4 | 2.3 | 81.8 | No |
| Walton | 2.3 | 7.6 | 4.6 | 2.6 | 23.6 | 10.2 | 0.7 | 15.0 | No |
| Washington | 8.3 | 25.5 | 13.2 | 13.7 | 11.0 | 4.2 | 0.3 | 88.2 | No |

Case rates nearly tripled in the period October 3 – November 14 for students in Florida compared to the period August 10 – October 3^7^ (4.5 per 1,000 in high school and 2.3 per 1,000 in elementary students to 12.5 and 7.4 per 1,000, respectively). Only two of Florida’s 67 counties had school case rates lower than the community case rate (Monroe and Walton), while 19 counties had higher pediatric case rates overall compared to school case rates (Table 2). Seven counties had more school cases reported than pediatric cases reported, which could be due to a number of high school students being over the age of 18 and thus not counted in the pediatric community rate, or do to the school reporting previous cases after-the-fact.

## Discussion

Case incidence varies significantly between school grade levels and between students and staff. Staff rates are higher than student rates in all school environments except high schools, and staff benefit most by mandatory-mask mandates. The rate of cases within schools is highly correlated with cases within a community, more than the size of the district by total enrollment. Percent enrollment in face-to-face instruction is a secondary influencer of case incidence rates in schools. In areas with higher pediatric community case rates compared to school case rates, districts may be under-reporting school case totals for students, or “disqualifying” student cases based on when a student tested and whether the case could be directly linked to the school environment. More research is needed to further understand the wealth of data available regarding COVID-19 incidence in Florida, and to develop proper mitigation strategies to confront this unprecedented challenge.

## Data Availability

All data is publicly available by request, and through our public data portal which may be accessed through our primary data site at www.TheCovidMonitor.com.

https://github.com/TheCovidMonitor/States/tree/main/Florida

https://experience.arcgis.com/experience/fb52d598982f41faac714b5ebe32e7d1

https://www.thecovidmonitor.com/

## Abbreviations

none

## Acknowledgements

The Covid Monitor provided data related to cases in K-12 school districts in Florida.

